# The effect of control strategies that reduce social mixing on outcomes of the COVID-19 epidemic in Wuhan, China

**DOI:** 10.1101/2020.03.09.20033050

**Authors:** Kiesha Prem, Yang Liu, Timothy W Russell, Adam J Kucharski, Rosalind M Eggo, Nicholas Davies, Centre for the Mathematical Modelling of Infectious Diseases COVID-19 Working Group, Mark Jit, Petra Klepac

## Abstract

**BACKGROUND:** In December 2019, a novel strain of SARS-CoV-2 emerged in Wuhan, China. Since then, the city of Wuhan has taken unprecedented measures and efforts in response to the outbreak.

**METHODS:** We quantified the effects of control measures on population contact patterns in Wuhan, China, to assess their effects on the progression of the outbreak. We included the latest estimates of epidemic parameters from a transmission model fitted to data on local and internationally exported cases from Wuhan in the age-structured epidemic framework. Further, we looked at the age-distribution of cases. Lastly, we simulated lifting of the control measures by allowing people to return to work in a phased-in way, and looked at the effects of returning to work at different stages of the underlying outbreak.

**FINDINGS:** Changes in mixing patterns may have contributed to reducing the number of infections in mid-2020 by 92% (interquartile range: 66–97%). There are benefits to sustaining these measures until April in terms of reducing the height of the peak, overall epidemic size in mid-2020 and probability that a second peak may occur after return to work. However, the modelled effects of social distancing measures vary by the duration of infectiousness and the role school children play in the epidemic.

**INTERPRETATION:** Restrictions on activities in Wuhan, if maintained until April, would likely contribute to the reduction and delay the epidemic size and peak, respectively. However, there are some limitations to the analysis, including large uncertainties around estimates of R0 and the duration of infectiousness.

**FUNDING:** Bill and Melinda Gates Foundation, National Institute for Health Research, Wellcome Trust, and Health Data Research UK.

## Introduction

The SARS-CoV-2, a novel coronavirus, emerged in the City of Wuhan, Hubei Province, China, in early December 2019.^1,2^ Since then, the local and national governments have taken unprecedented measures in response to the COVID-19 outbreak caused by SARS-CoV-2.^3^ Exit screening of passengers was shortly followed by travel restrictions in Wuhan on 23^rd^ January 2020, halting all means of unauthorised travel into and out of the city. Similar control measures were extended to the entire province of Hubei by 26^th^ January 2020.^3^ Non-pharmaceutical social distancing interventions such as extended school closure and workplace distancing were introduced to reduce the impact of the COVID-19 outbreak in Wuhan.^4^ Within the city, schools remained closed; Lunar New Year holidays were extended so that people stayed away from their workplaces; the local government promoted social distancing and encouraged residents to avoid crowded places. These measures are known to greatly changed the age-specific mixing patterns within the population in previous outbreak response efforts for other respiratory infectious diseases.^5,6^ While travel restrictions undoubtedly had a role in reducing the exportations of infections outside of Wuhan, and delayed the onset of outbreaks in other regions,^7,8^ changes in the mixing patterns affected the trajectory of the outbreak within Wuhan itself. In order to estimate the effects of social distancing measures on the progression of the COVID-19 epidemic, we look at Wuhan, hoping to provide some insights for the rest of the world.

Person-to-person transmission is mostly driven by “who interacts with whom”,^9,10^ which can vary by age and location of the contact i.e., school, work, home, and community. Under the context of a large-scale on-going outbreak, contact patterns would drastically shift from their baseline conditions. In the COVID-19 outbreak in Wuhan, social distancing measures including but not limited to school and workplace closures and health promotions that encourage the general public to avoid crowded places are designed to drastically shift social mixing patterns and are often used in epidemic settings.^4^ While contact patterns can be inferred from reported social contact data that include the information in which setting the contact took place, such studies are often focused on high-income countries,^11^ or particular high-density areas.^12^ Prem and colleagues^13^ address that limitation by quantifying contact patterns in the home, school, work and other locations across a range of countries based on available information from household-level data and local population demographic structure.

To examine how these changes in population mixing have affected the outbreak progression in Wuhan, we used synthetic location-specific contact patterns in Wuhan and adapted it in the presence of school closures, extended workplace closures, reduction in mixing in the general community. Using these matrices and the latest estimates of the epidemiological parameters of the Wuhan outbreak,^14–16^ we simulated the ongoing trajectory of an outbreak in Wuhan using an age-structured susceptible-exposed-infectious-removed (SEIR) model^17,18^ for several social distancing measures.

## Methods

### SEIR model

We simulated the outbreak in Wuhan using a deterministic stage-structured SEIR model over a six month period, during which the modelled outbreak peters out. An implication of this approach is that all demographic changes in the population (i.e., births, deaths, and ageing) are ignored.

We divide the population according to the infection status into susceptible (*S*), exposed (*E*), infected (*I*), and removed (*R*) individuals, and according to age into five-year bands until age 75 and a single category aged 80+ (resulting in the total of 16 age categories, *n*). Susceptible individuals may acquire the infection at a given rate when they come into contact with an infectious person, and enter the exposed disease state before they become infectious and later either recover or die. We assume Wuhan to be a close system with a constant population size of 11 million (i.e., *N* = *S* + *E* + *I* + *R* = 11 million) throughout the course of this epidemic. We considered the SEIR model presented in **Figure 1**. The age-specific mixing patterns of individuals in age group *i* alter their likelihood of being exposed to the virus given a certain number of infectives in the population. In addition, we incorporated contributions of asymptomatic and subclinical cases, however, the question of whether they are able to transmit infection is still not resolved at the time of writing, although current evidence suggests that they are likely to.^19^ We further considered a scenario where we assumed that younger individuals are more likely to be asymptomatic (or subclinical) and less infectious than older individuals.^20,21^

**Figure 1.**
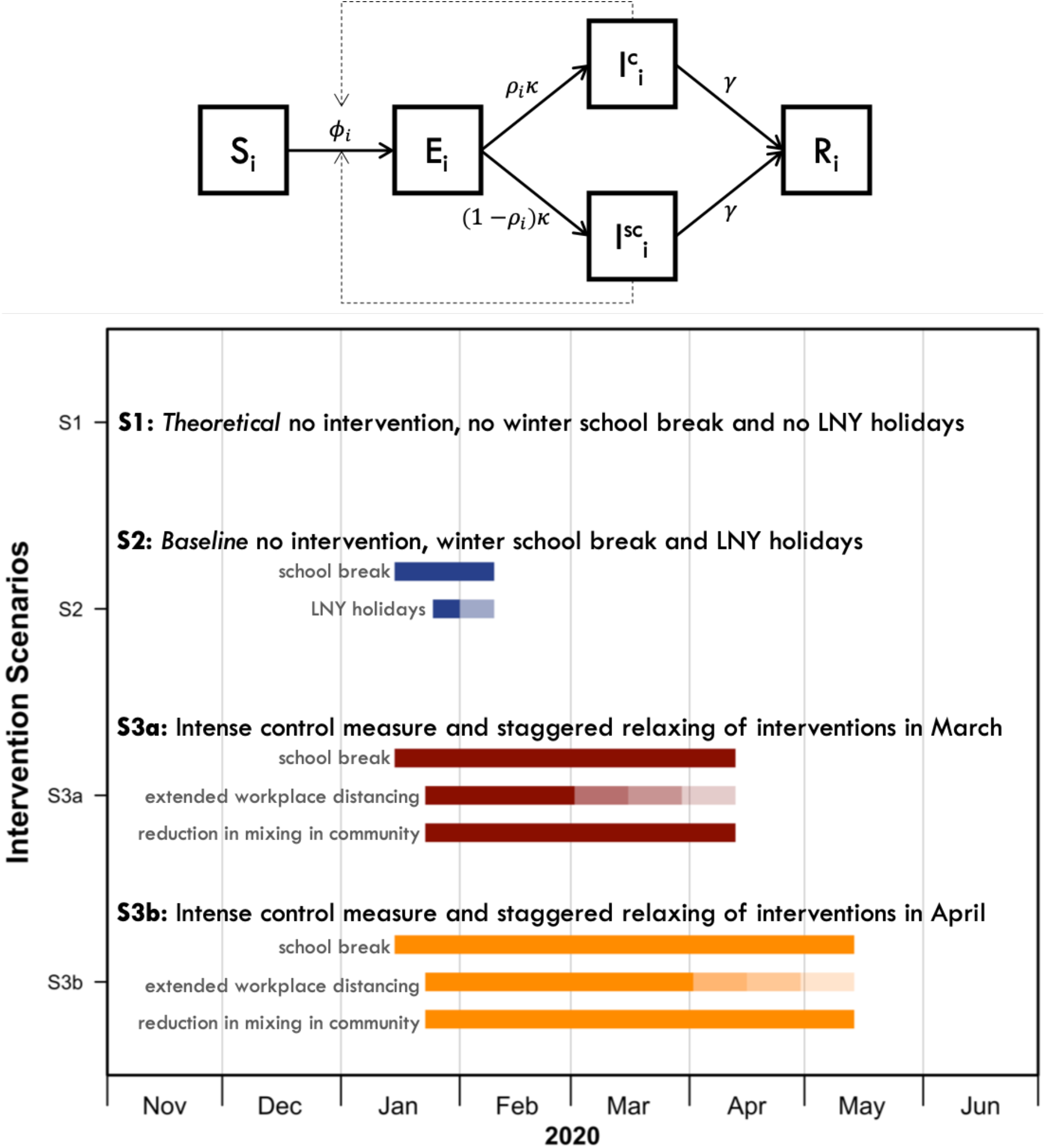
Age-structured SEIR model and details of the modelled social distancing interventions. The age-specific mixing patterns of individuals in age group *i*alter their likelihood of being exposed to the virus given a certain number of infectives in the population. Younger individuals are more likely to be asymptomatic and less infectious, i.e. subclinical (SC). When *ρ*_*i*_ = 0 for all *i*, the model simplifies to a standard SEIR. The force of infection *ϕ*_*i,t*_ is given by 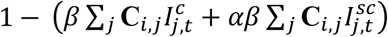.

For a given age group *i*, epidemic transitions can be described by:

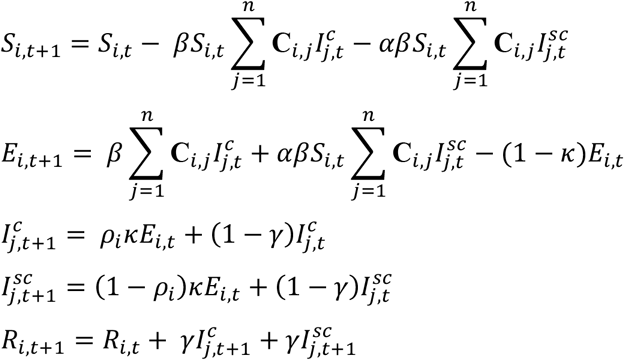

Where *β* is the transmission rate (scaled to the right value of *R*_0_), **C**_*i,j*_ describe the contacts of age group *j* made by age group *i, κ* = 1 − exp (−1/*d*_*L*_) is the daily probability of exposed individual becoming infectious (with *d*_*L*_ being the average incubation period), and *γ* = 1 − exp (−1/*d*_*I*_) is the daily probability that an infected individual recovers when the average duration of infection is *d*_*I*_. We further incorporated contributions of asymptomatic and subclinical cases, 1 − *ρ*_*i*_ denotes the probability of an infected cases being asymptomatic or subclinical. We assumed that younger individuals are more likely to be asymptomatic (or subclinical) and less infectious (proportion of infectiousness compared to *I*^*c*^, *α*).

Using parameters from the literature as presented in **Table 1**, we simulated the outbreak. In particular, we assumed the average incubation period and average infectious period to be 6.4 days^15^ and 3 or 7 days, respectively. Each simulation started with 200 or 2000 infectious individuals *I*_0_,^16^ with the rest of the population being in the susceptible state. We explored the uncertainty in the model by drawing *R*_0_ values uniformly from the 95% confidence interval from the posterior of the *R*_0_ distribution from the semi-mechanistic model by Kucharski and colleagues^14^ (**Fig S3**).

**Table 1.** Parameters of the SEIR model.

| Parameter | Values | References |
| --- | --- | --- |
| Basic reproduction number, $R_0$ | 2.2 (1.6–3.0)<sup>1</sup> | Kucharski and colleagues<sup>14</sup> |
| Average incubation period, $d_L$ | 6.4 days | Backer and colleagues<sup>15</sup> |
| Average duration of infection, $d_I$ | 3 or 7 days | (assumed) |
| Initial number of infected, $I_0$ | 200 or 2000 | Abbott and colleagues<sup>16</sup> |
| Pr(infected case is clinical), $\rho_i$ | 0 or 0.4, for $i \leq 4$ | Bi and colleagues<sup>20</sup> |
| | 0 or 0.8, for $i > 4$ | Davies and colleagues<sup>21</sup> |
| Pr(infection acquired from subclinical), $\alpha$ | 0.25 | Liu and colleagues<sup>19</sup> |

<sup>1</sup>Median and interquartile range are presented.

### Social mixing and interventions

Social mixing patterns vary across locations—households, workplaces, schools, and other locations. Therefore, we use the method set out in Prem and colleagues^13^ which accounts for these differences, and obtain the location-specific contact matrices **C** for different scenarios. In a normal setting, contacts made at all of these locations contribute to the overall mixing pattern in a population, so we sum the contacts across the different locations to obtain our baseline contact pattern in the population before the outbreak (**Fig 2, Fig S1** and **Fig S2**). In an outbreak setting, different intervention strategies are aimed at reducing social mixing in different contexts in order to lower the overall transmission in the population. To simulate the effects of interventions aimed at reducing social mixing, we create synthetic contact matrices for each intervention scenario from these building block matrices.

**Figure 2.**
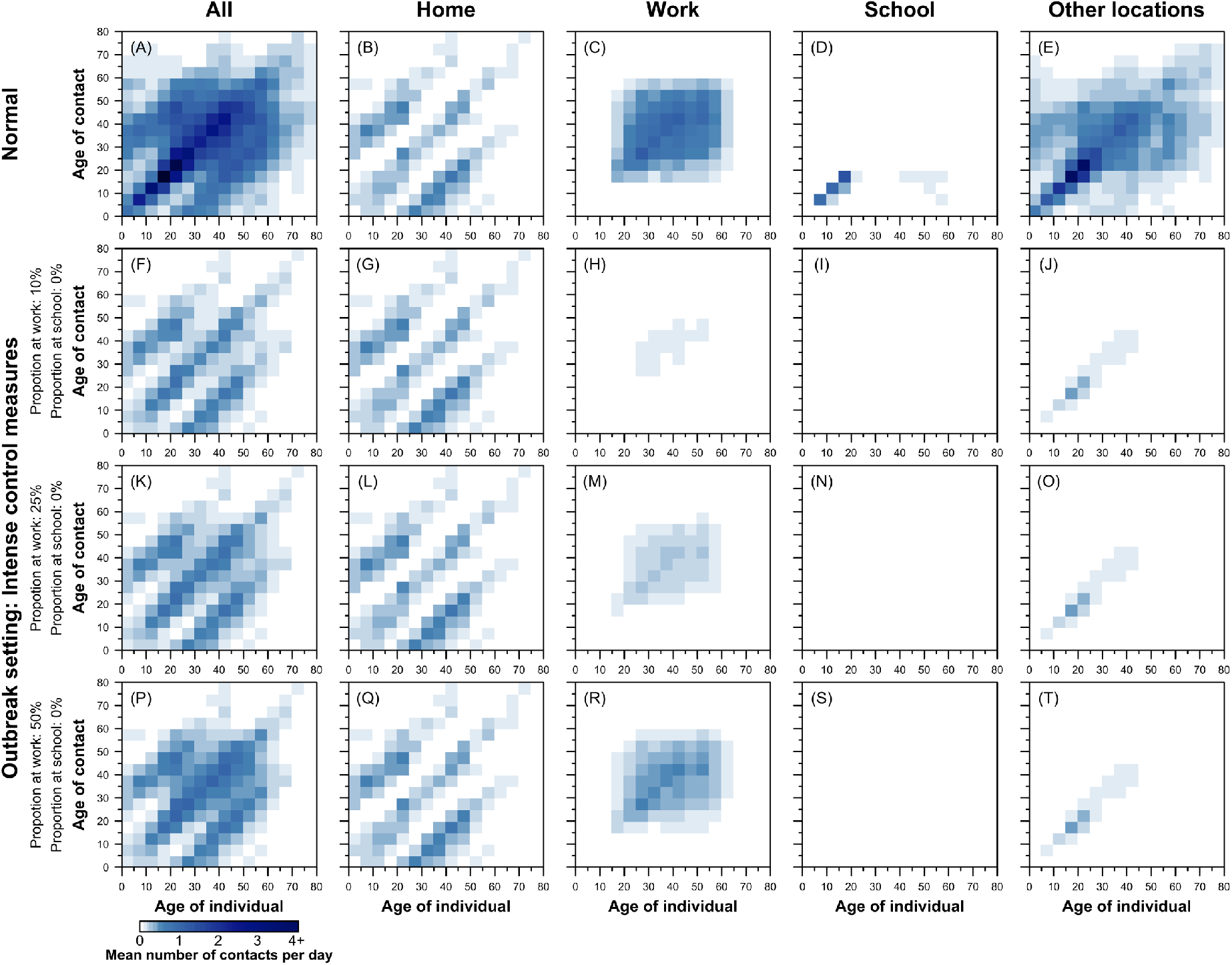
Synthetic age- and location-specific contact matrices for China under various social distancing scenarios during the intense control period. The synthetic age-specific contact patterns across all locations, at home, at the workplace, in school, and at other locations during normal circumstances (i.e. under no intervention) are presented in panels A–E. The age- and location-specific contact matrices under the various social distancing interventions are presented in the panels F–T. Darker colour intensities indicate higher proclivity of making the age-specific contact.

We consider the following three scenarios: (1) *theoretical*: assume no change to social mixing patterns at all location types, no school term break and no Lunar New Year holidays; (2) *no interventions, winter school break in Wuhan, and Lunar New Year holidays*: assume no social distancing control measures, school-going individuals do not have any contacts at school because of school holidays from 15^th^ January–10^th^ February 2020 and 10% and later 75% of workforce will be working during the holidays from 25^th^–31^st^ January 2020 and from 1^st^ –10^th^ February 2020, respectively; (3) *intense control measures in Wuhan to contain the outbreak*: assume school closure and about 10% of workforce—for example, healthcare personnel, police, other essential government staff—will be working even during the control measures (**Fig 2** and **Fig S4**). For the third scenario, we further modelled the impact of whether the intense control measures end in beginning of March or April, and we allowed for a staggered return to work while the school remains closed, i.e. 25% of workforce will be working in weeks one and two (**Fig 2** second row), 50% of workforce will be working in weeks three and four to work (**Fig 2** third row), and 100% of workforce will be working and school resumes (**Fig 2** fourth row). ^3,22,23^

## Results

Our simulations show that the control measures aimed at reducing the social mixing in the population can be effective in reducing the magnitude of the peak of the outbreak. **Figure 3** shows the effects of different control measures among individuals aged 55–60 and 10–15 years old. The standard school winter break and holidays for the Lunar New Year would have had little effect on progression of the outbreak had the school and workplace re-opened as normal.

**Figure 3.**
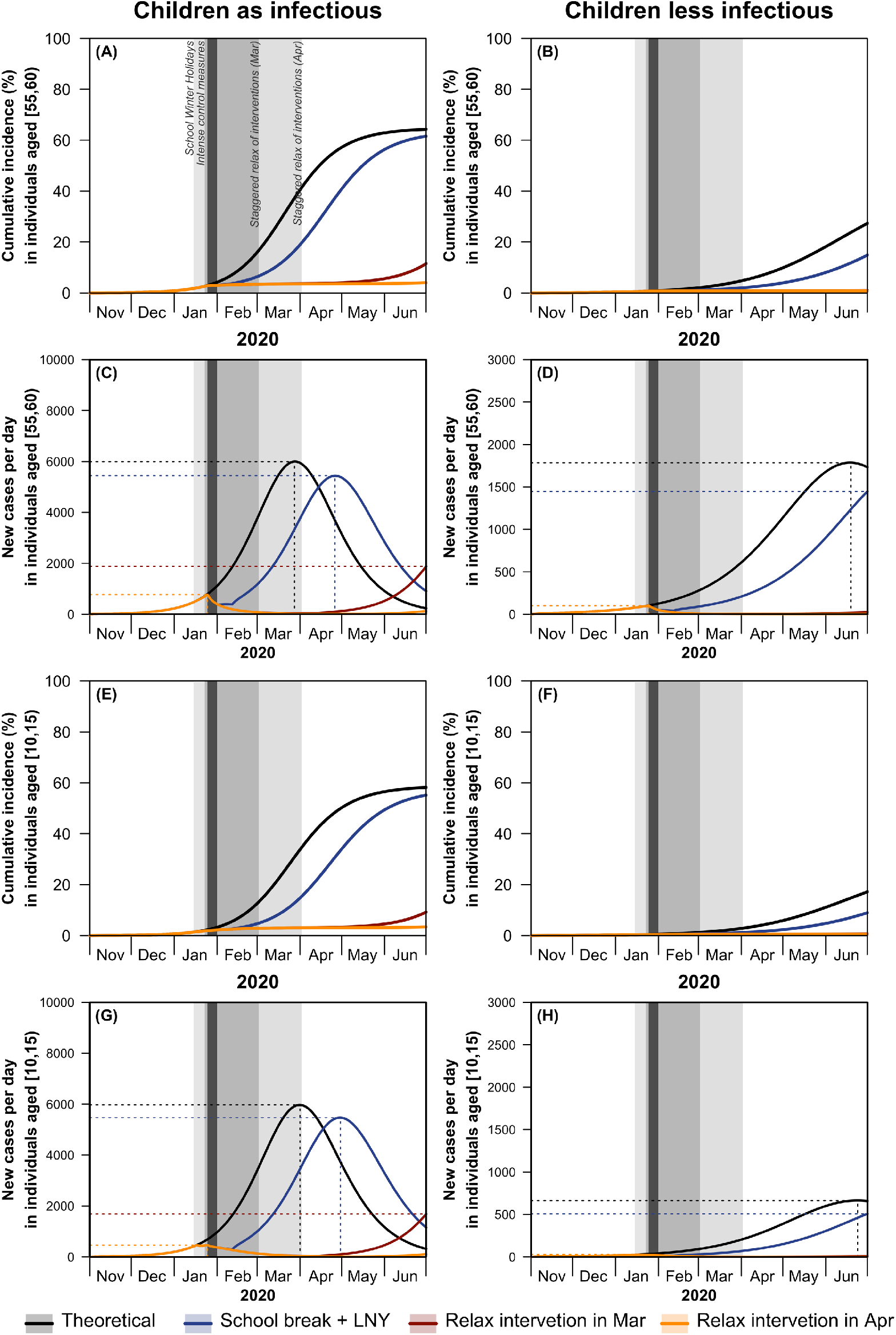
Effects of different intervention strategies on the cumulative incidence and new cases per day among individuals aged 55–60 (A to D) and 10–15 (E to H) from late 2019 to mid-2020. Under two scenarios children being equally infectious and children being less infectious, the effects of social distancing measures were investigated. Theoretical no intervention (black line), school break and LNY (blue line) and intense control measures that are relaxed in a staggered fashion at the beginning of March (red line), and intense control measures that are relaxed in a staggered fashion at the beginning of April (orange line). Shading indicates the timing of the school holidays, Lunar New Year (dark vertical line), intense control measures (dark grey), and staggered return to work followed by school opening (lighter grey).

We presented the median cumulative incidence (**Fig 4A**), incident case per day (**Fig 4B**) and age-specific incidence per day (**Fig 4C–G**) of the 200 simulated outbreaks. The 25^th^ and 75^th^ percentile outbreaks are represented by the shaded area in the cumulative incidence. The intense control measures of prolonged school closure and work holidays reduced the final size (**Fig 4A**) and peak incidence, while also delaying the peak of the outbreak (**Fig 4B**). Our model suggests that the effects of these social distancing strategies vary across age categories, the reduction in incidence is highest among school children and older individuals and lowest among working-aged adults (**Fig 4C–G** and **Fig 5**).

**Figure 4.**
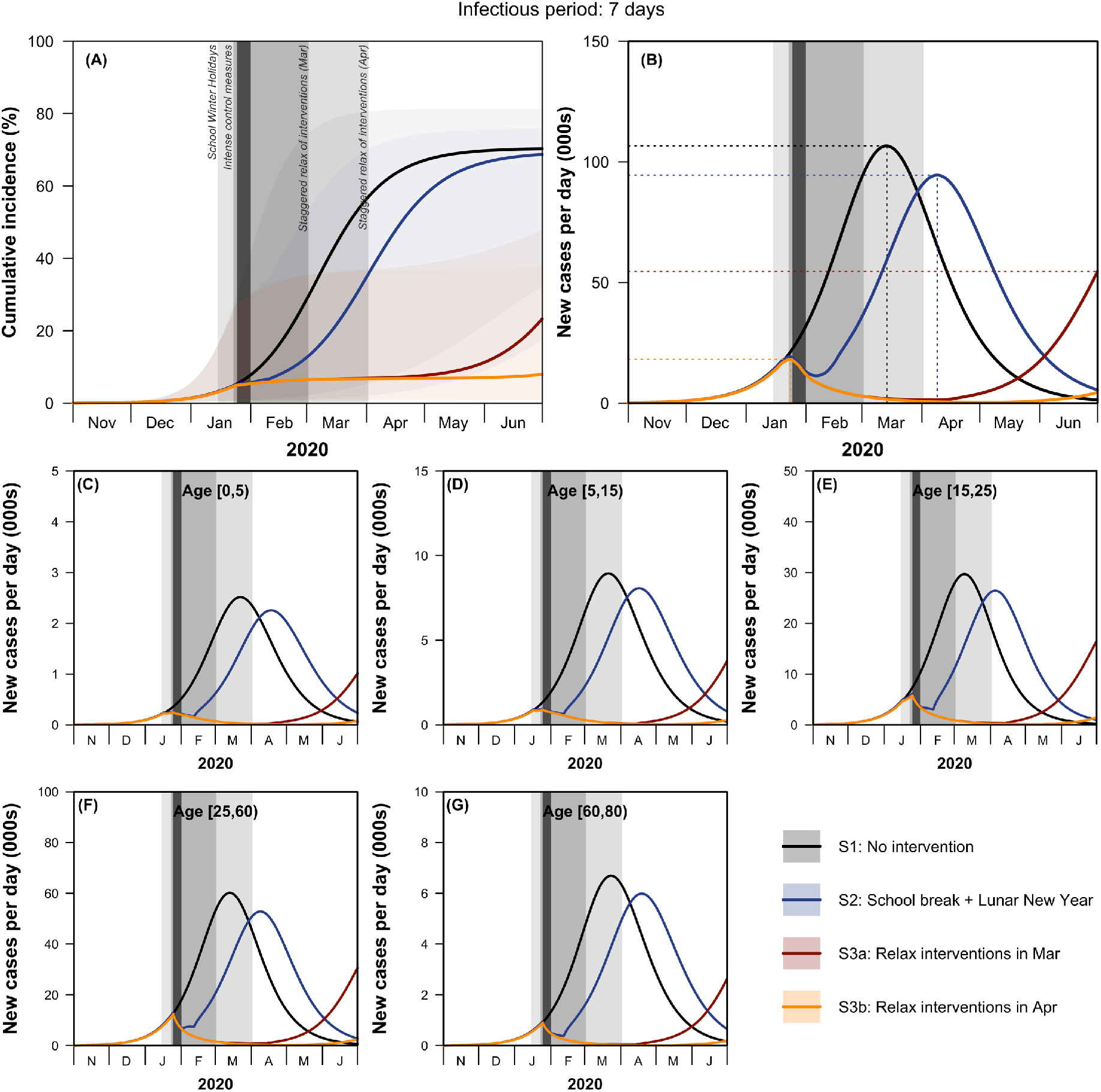
Effects of the different social distancing measures on the cumulative incidence (A) and new cases per day (B), and age-specific incidence per day (C to G) from late 2019 to mid-2020. The median cumulative incidence, incident cases per day and age-specific incidence per day are represented as solid lines. The 25^th^ and 75^th^ percentile outbreaks are represented by the shaded area in the cumulative incidence. Theoretical no intervention (black line), school break and Lunar New Year (blue line) and intense control measures that are relaxed in a staggered fashion at the beginning of March (red line), and intense control measured that are relaxed in a staggered fashion at the beginning of April (orange line). Shading indicates the timing of the school holidays, Lunar New Year weekend (dark vertical line), intense control measures (dark grey), and staggered return to work followed by school opening (lighter grey).

**Figure 5.**
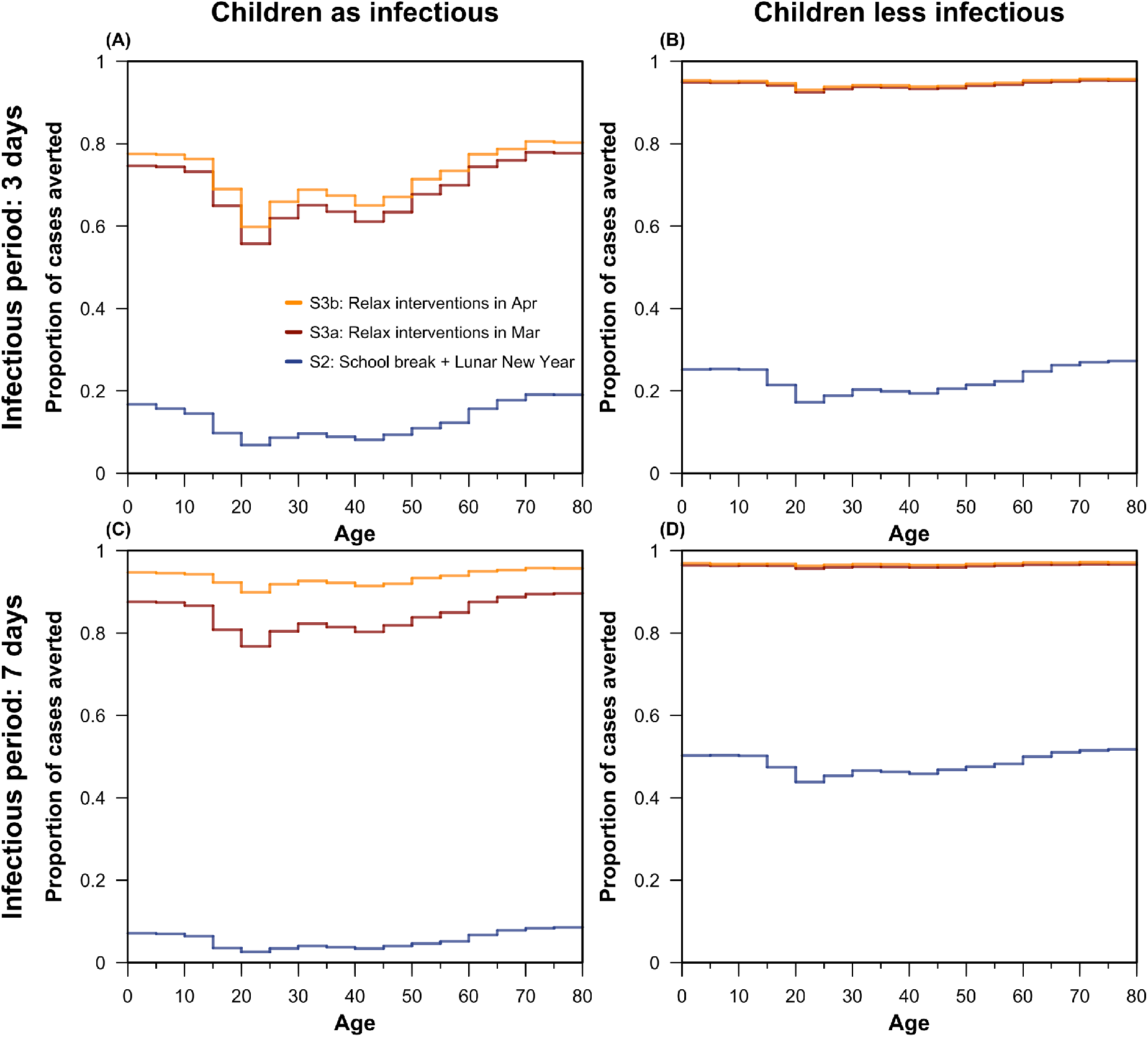
The modelled proportion of number of infections averted in mid-2020 by age for the different social distancing measures, assuming the duration of infectiousness to be (A and B) 3 days and (C and D) 7 days. The additional proportions of cases averted (compared to no intervention) are presented across age and by the different social distancing measures: school break and Lunar New Year (blue area), intense control measures that are relaxed in a staggered fashion at the beginning of March (red area), and intense control measures that are relaxed in a staggered fashion at the beginning of April (orange area).

These measures were most effective if staggered return to work was at the beginning of April; this reduced the overall number of infections in mid-2020 size by more than 92% (interquartile range: 66– 97%) (**Fig 5** and **Fig S4**) should the disease have a longer duration of infectiousness, and greatly reduced the magnitude of the peak incidence across all age categories (**Fig 4C–E**) which can have further beneficial impact by relieving the pressure on the healthcare system. However, premature return to work can result in an increase in incidence even if it originally seemed that the epidemic has started to turn over (second wave, **Fig 4B** peak of the orange line followed by the sharp increase in incidence in red line). Uncertainty in *R*_0_ values has a large impact on the timing of the peak and the final size of the outbreak (**Fig 4A**).

The modelled effects of intense control measures of prolonged school closure and work holidays vary by the duration of infectiousness. If the disease had a short infectious period (3 days), then the model suggests that relaxing the social distancing interventions in March (**Fig 5A**) could avert approximately half of cases in school children and older individuals. More than half of the cases could be averted should the disease have a longer duration of infectiousness (for e.g., 7 days as presented in **Fig 5C**)— social distancing interventions would need to be relaxed a month later (in April) in order to observe a larger effect. If children were less infectious as described in M2, lifting the social distancing interventions in April instead of March could have less additional health benefits (**Fig 5B** and **5D**).

## Discussion

SARS-CoV-2, a contact-transmissible infectious disease, percolates through a population through direct contact between infected and susceptible individuals.^1,9,10^ Outbreak control measures aimed at reducing the amount of mixing in the population have the potential to reduce the final size of the epidemic and the magnitude of the peak. To evaluate the effect of location-specific social distancing measures such as extended school closure and interventions around workplaces on the magnitude of the peak and the final size, we accounted for these heterogeneities in contact networks in our model. Here, we simulated outbreaks and modelled the interventions by scaling down the appropriate component of the contact mixing matrices for China.

Mathematical models can help us understand how SARS-CoV-2 would spread across the population and inform control measures that may mitigate future transmission.^24,25^ Here, we simulated the trajectory of the ongoing outbreak of COVID-19 in Wuhan, using an age-structured SEIR model.^17,18^ Because individuals’ mixing patterns are non-random, they influence the transmission dynamics of the disease.^11^ Models assessing the effectiveness of social distancing interventions, such as school closure, need to account for social structures and heterogeneities in mixing of individuals.^26–30^ In our model, we incorporated changes to age- and location-specific social mixing patterns to estimate the effects of location-specific social distancing interventions in curtailing the spread of the outbreak. Consequently, if these restrictions are lifted prematurely while there are still enough susceptibles to keep the *R*_*e*_ >1once contacts increase, the number of infections would increase. Realistically, interventions are lifted slowly, partly as an attempt to avoid a sharp increase in infection, but also for logistical and practical reasons. Therefore, lifting the interventions was simulated in a staggered fashion, whereby the interventions were relaxed bit-by-bit (**Figures 3, 4** and **5**).

Current evidence of the effects of various social distancing measures on containing the outbreak are limited and little is known about the behavioural changes of individuals over a period of time either during an outbreak, or otherwise. Therefore, to model the effects of the social distancing measures implemented in Wuhan, we assumed what effect certain types of social distancing has on age- and location-specific contact rates.

Much remains to be discovered about the true age-specific susceptibility and transmissibility of COVID-19. Therefore, we assumed no heterogeneity in susceptibility between children. Furthermore, for simplicity we assumed children and adults were equally transmissible, other than the differences in their contact rates. Similar to a flu-like pathogen, the model suggests that interactions between school children and the older individuals in the population have important public health implications, as children may have high infection rates while the elderly are more vulnerable to severe infections with potentially fatal outcome.^31,32^ However, unlike models built for pandemic or seasonal flu, we do take into account the lack of population immunity to SARS-CoV-2.

Extreme social distancing measures, including school closures, workplace closures, and avoiding any public gatherings all at once, can push the transmission to households leading to increased clustering in household cases.^5^ As households are not explicitly included in the model, we do not consider heterogeneity and clustering of household transmission. Distinguishing between repeated and new contacts is important for disease propagation in contact network models,^33,34^ more sophisticated methods accounting for temporal presence within the household^35^ would be needed to characterise higher degrees of contact.

A key parameter is the basic reproduction number (*R*_0_) which determines how fast SARS-CoV-2 spreads through the population during the early stages of the outbreak. This is an inherently difficult parameter to estimate, since the true number of cases that can transmit infection at a given time is unknown (reported cases are likely to be just a small fraction of these), and likely varies over time (due to different interventions being introduced and population behaviour changing in response to the epidemic). In our analysis, we used an existing model that inferred time-dependent *R*_*e*_ based on the growth of reported cases in Wuhan as well as number of exported cases outside China originating from Wuhan.^14^

Social distancing and travel restrictions combined have clearly aided the lowering of the transmission of COVID-19 over the course of the ongoing outbreak in Wuhan.^8,36,37^ Evidence for this drop in transmission can be gleaned from the time-varying estimates of the reproductive number,^14^ or, even more simply, observing that the turnover of the epidemic has occurred far before the depletion of the susceptibles indicates the effects of the implemented measures. Insofar as social distancing alone is responsible for the drop is extremely difficult to quantify, especially during the ongoing epidemic. Therefore we took a broad-view of the question, making assumptions about the result of certain forms of social distancing and measuring the effects somewhat qualitatively. However, it is clear that to a greater or lesser extent, social distancing has resulted in both a shorter epidemic and one with a lower peak. Given what is known about the transmissibility and (the relatively long) incubation period of COVID-19,^1,15^ the efficacy of social distancing in reducing these important attributes of any epidemic are no surprise.

## Conclusion

Non-pharmaceutical interventions based on sustained social distancing have a strong potential to reduce the magnitude of the peak and lead to overall smaller number of cases. Lowering and flattening of the epidemic peak is particularly important, as it reduces the acute pressure on the health system. Premature and sudden lifting of interventions could lead to a secondary peak that can be flattened by relaxing the interventions gradually.

## Data Availability

All data and code will be made available.

https://github.com/kieshaprem/ncov-agestructured-seir

## Contributors

PK, YL, MJ, and KP conceived the study. KP, YL, and PK designed and programmed the model, and KP made the figures. TWR, AJK, RME, and ND consulted on the analyses. All authors interpreted the results, contributed to writing the Article, and approved the final version for submission.

## Acknowledgment

KP, YL, MJ, and PK were funded by the Bill & Melinda Gates Foundation (grant number INV-003174), YL and MJ were funded by the National Institute for Health Research (NIHR) (16/137/109), TWR and AJK were funded by the Wellcome Trust (grant number 206250/Z/17/Z), RME was funded by HDR UK (grant number MR/S003975/1), and ND was funded by NIHR (HPRU-2012-10096).This research was partly funded by the National Institute for Health Research (NIHR) (16/137/109) using UK aid from the UK Government to support global health research. The views expressed in this publication are those of the author(s) and not necessarily those of the NIHR or the UK Department of Health and Social Care. We would like to acknowledge (in a randomised order) the other members of the London School of Hygiene & Tropical Medicine COVID-19 modelling group, who contributed to this work: Stefan Flasche, Samuel Clifford, Carl A B Pearson, James D Munday, Sam Abbott, Hamish Gibbs, Alicia Rosello, Billy J Quilty, Thibaut Jombart, Fiona Sun, Charlie Diamond, Amy Gimma, Kevin van Zandvoort, Sebastian Funk, Christopher I Jarvis, W John Edmunds, Nikos I Bosse, and Joel Hellewell. Their funding sources are as follows: Stefan Flasche and Sam Clifford (Sir Henry Dale Fellowship [grant number 208812/Z/17/Z]); Billy J Quilty, Fiona Sun, and Charlie Diamond (NIHR [grant number 16/137/109]); Joel Hellewell, Sam Abbott, James D Munday, and Sebastian Funk (Wellcome Trust [grant number 210758/Z/18/Z]); Amy Gimma and Christopher I Jarvis (Global Challenges Research Fund [grant number ES/P010873/1]); Hamish Gibbs (Department of Health and Social Care [grant number ITCRZ 03010]); Alicia Rosello (NIHR [grant number PR-OD-1017-20002]); Thibaut Jombart (RCUK/ESRC [grant number ES/P010873/1], UK PH RST, NIHR HPRU Modelling Methodology); Kevin van Zandvoort (Elrha’s Research for Health in Humanitarian Crises (R2HC) Programme, UK Government (DFID), Wellcome Trust, NIHR).

## References

1. Li Q, Guan X, Wu P, et al. Early Transmission Dynamics in Wuhan, China, of Novel Coronavirus–Infected Pneumonia. N Engl J Med 2020; published online Jan 29. DOI:10.1056/nejmoa2001316.

2. Zhu N, Zhang D, Wang W, et al. A Novel Coronavirus from Patients with Pneumonia in China, 2019. N Engl J Med 2020; published online Jan 24. DOI:10.1056/nejmoa2001017.

3. Chen S, Yang J, Yang W, Wang C, Bärnighausen T. COVID-19 control in China during mass population movements at New Year. Lancet (London, England) 2020; published online Feb 24. DOI:10.1016/S0140-6736(20)30421-9.

4. Fong MW, Gao H, Wong JY, et al. Nonpharmaceutical Measures for Pandemic Influenza in Nonhealthcare Settings—Social Distancing Measures. Emerg Infect Dis 2020; 26. DOI:10.3201/eid2605.190995.

5. Hens N, Ayele GM, Goeyvaerts N, et al. Estimating the impact of school closure on social mixing behaviour and the transmission of close contact infections in eight European countries. BMC Infect Dis 2009; 9: 187.

6. Ahmed F, Zviedrite N, Uzicanin A. Effectiveness of workplace social distancing measures in reducing influenza transmission: A systematic review. BMC Public Health. 2018; 18. DOI:10.1186/s12889-018-5446-1.

7. Quilty BJ, Clifford S, Flasche S, Eggo RM. Effectiveness of airport screening at detecting travellers infected with novel coronavirus (2019-nCoV). Eurosurveillance 2020; 25: 2000080.

8. Tian H, Li Y, Liu Y, et al. Early evaluation of the Wuhan City travel restrictions in response to the 2019 novel coronavirus outbreak. medRxiv 2020; : 2020.01.30.20019844.

9. Riou J, Althaus CL. Pattern of early human-to-human transmission of Wuhan 2019 novel coronavirus (2019-nCoV), December 2019 to January 2020. Euro Surveill 2020; 25. DOI:10.2807/1560-7917.ES.2020.25.4.2000058.

10. Chan JFW, Yuan S, Kok KH, et al. A familial cluster of pneumonia associated with the 2019 novel coronavirus indicating person-to-person transmission: a study of a family cluster. Lancet 2020; 395: 514–23.

11. Mossong J, Hens N, Jit M, et al. Social Contacts and Mixing Patterns Relevant to the Spread of Infectious Diseases. PLoS Med 2008; 5: e74.

12. Zhang J, Klepac P, Read JM, et al. Patterns of human social contact and contact with animals in Shanghai, China. Sci Rep 2019; 9: 1–11.

13. Prem K, Cook AR, Jit M. Projecting social contact matrices in 152 countries using contact surveys and demographic data. PLoS Comput Biol 2017; 13. DOI:10.1371/journal.pcbi.1005697.

14. Kucharski AJ, Russell TW, Diamond C, group C nCoV working, Funk S, Eggo RM. Early dynamics of transmission and control of 2019-nCoV: a mathematical modelling study. medRxiv 2020; : 2020.01.31.20019901.

15. Backer JA, Klinkenberg D, Wallinga J. Incubation period of 2019 novel coronavirus (2019-nCoV) infections among travellers from Wuhan, China, 20–28 January 2020. Eurosurveillance 2020; 25: 2000062.

16. Abbott S, Hellewell J, Munday J, Funk S. The transmissibility of novel Coronavirus in the early stages of the 2019-20 outbreak in Wuhan: Exploring initial point-source exposure sizes and durations using scenario analysis. Wellcome Open Res 2020; 5: 17.

17. Klepac P, Pomeroy LW, Bjørnstad ON, Kuiken T, Osterhaus ADME, Rijks JM. Stage-structured transmission of phocine distemper virus in the Dutch 2002 outbreak. Proc R Soc B Biol Sci 2009; 276: 2469–76.

18. Klepac P, Caswell H. The stage-structured epidemic: Linking disease and demography with a multi-state matrix approach model. Theor Ecol 2011; 4: 301–19.

19. Liu Y, Funk S, Flasche S. The Contribution of Pre-symptomatic Transmission to the COVID-19 Outbreak | CMMID Repository. https://cmmid.github.io/topics/covid19/control-measures/pre-symptomatic-transmission.html (accessed March 2, 2020).

20. Bi Q, Wu Y, Mei S, et al. Epidemiology and Transmission of COVID-19 in Shenzhen China: Analysis of 391 cases and 1,286 of their close contacts. medRxiv 2020; : 2020.03.03.20028423.

21. Davies N. nicholasdavies/ncov-age-dist. https://github.com/nicholasdavies/ncov-age-dist (accessed March 6, 2020).

22. Zhengzhou Daily. Zhezhou City Strategy for Resuming Work and Production Among Business Enterprises. 2020; published online Feb. http://m.xinhuanet.com/ha/2020-02/13/c_1125567288.htm.

23. Sohu News. Academic Calendar for Elementary and Middle School in Wuhan. 2019. https://www.sohu.com/a/325806609_500181.

24. Riley S, Fraser C, Donnelly CA, et al. Transmission dynamics of the etiological agent of SARS in Hong Kong: Impact of public health interventions. Science (80-) 2003; 300: 1961–6.

25. Ferguson NM, Keeling MJ, Edmunds WJ, et al. Planning for smallpox outbreaks. Nature 2003; 425: 681–5.

26. Wallinga J, Teunis P, Kretzschmar M. Using Data on Social Contacts to Estimate Age-specific Transmission Parameters for Respiratory-spread Infectious Agents. Am J Epidemiol 2006; 164: 936–44.

27. Read JM, Keeling MJ. Disease evolution on networks: the role of contact structure. Proc R Soc London B Biol Sci 2003; 270: 699–708.

28. Hilton J, Keeling MJ. Incorporating household structure and demography into models of endemic disease. J R Soc Interface 2019; 16. DOI:10.1098/rsif.2019.0317.

29. Wallinga J, Edmunds WJJ, Kretzschmar M. Perspective: human contact patterns and the spread of airborne infectious diseases. Trends Microbiol 1999; 7: 372–7.

30. Edmunds WJ, O’callaghan CJ, Nokes DJ. Who mixes with whom? A method to determine the contact patterns of adults that may lead to the spread of airborne infections. Proc R Soc B Biol Sci 1997; 264: 949–57.

31. Cate TR. Clinical manifestations and consequences of influenza. Am J Med 1987; 82: 15–9.

32. Falsey AR, Erdman D, Anderson LJ, Walsh EE. Human Metapneumovirus Infections in Young and Elderly Adults. J Infect Dis 2003; 187: 785–90.

33. Eubank S, Guclu H, Anil Kumar Vs, et al. Modelling disease outbreaks in realistic urban social networks. Nature 2004; 429: 180–4.

34. Stehlé J, Voirin N, Barrat A, et al. Simulation of an SEIR infectious disease model on the dynamic contact network of conference attendees. BMC Med 2011; 9: 87.

35. Potter GE, Handcock MS, Longini IM, Halloran ME. Estimating Within-Household Contact Networks from Egocentric Data. Ann Appl Stat 2011; 5: 1816–38.

36. Brooks SK, Webster RK, Smith LE, et al. The psychological impact of quarantine and how to reduce it: rapid review of the evidence. Lancet 2020; published online Feb 26. DOI:10.1016/S0140-6736(20)30460-8.

37. Wu JT, Leung K, Leung GM. Nowcasting and forecasting the potential domestic and international spread of the 2019-nCoV outbreak originating in Wuhan, China: a modelling study. Lancet 2020; 395: 689–97.

